# HG-DCM: History Guided Deep Compartmental Model for Early Stage Pandemic Forecasting

**DOI:** 10.1101/2024.11.18.24317469

**Authors:** Ziming Wei, Michael Lingzhi Li

## Abstract

We introduce the History-Guided Deep Compartmental Model (HG-DCM), a novel framework for early-stage pandemic forecasting that synergizes deep learning with compartmental modeling to harness the strengths of both interpretability and predictive capacity. HG-DCM employs a Residual Convolutional Neural Network (RCNN) to learn temporal patterns from historical and current pandemic data while incorporating epidemiological and demographic metadata to infer interpretable parameters for a compartmental model to forecast future pandemic growth. Experimental results on early-stage COVID-19 and Monkeypox forecasting tasks demonstrate that HG-DCM outperforms both standard compartmental models (e.g., DELPHI) and standalone deep neural networks (e.g., GRU) in predictive accuracy and stability, particularly with limited data. By effectively integrating historical pandemic insights, HG-DCM offers a scalable approach for interpretable and accurate forecasting, laying the groundwork for future real-time pandemic modeling applications.

## 1 Introduction

Pandemics have historically caused catastrophic losses, from the Bubonic Plague in the 14th century [24] to the smallpox outbreak in the 18th century [11], and most recently, the COVID-19 pandemic in 2020 [13]. Despite significant advances in medical science, technology, and epidemiology, COVID-19 alone resulted in millions of deaths worldwide from 2020 to 2023. Accurate early-stage estimation of pandemic severity remains a crucial topic - Studies suggest that with improved forecasting and prompt interventions, early pandemic mortality could be reduced by as much as 90% [28, 19]. Yet accurate early-warning prediction is fundamentally challenging, with the lack of high-quality data being a major challenge. Mispredictions of pandemic severity lead to significant consequences: Underestimating an outbreak risks overwhelming healthcare systems and delaying crucial interventions, thereby increasing mortality and transmission rates. Conversely, overestimations can lead to inefficient use of resources and societal disruptions, including panic buying [15, 3] and social unrest [1, 29].

A significant number of current pandemic forecasting models are compartmental models, in which the incidence of each location is fit separately and completely relies on data specific to the current outbreak. The limited data source of compartmental models leads to unsatisfactory performance on early pandemic forecasting tasks. Past pandemics can provide significant information on the likely severity of the current pandemic at the early stage, but compartmental models lack the ability to integrate past pandemic information into forecasting. The wealth of historical pandemic data, which, though costly in terms of human lives, remains underutilized and represents a missed opportunity to enhance predictive accuracy. Therefore, in this study, we present the History-Guided Deep Compartmental Model (HG-DCM), which leverages historical data and meta-data to enhance forecasting accuracy by incorporating insights from previous pandemics and early-stage pandemic meta-data.

HG-DCM combines a residual convolutional neural network [12] with a novel compartmental model named DELPHI [19] to create a powerful tool for early pandemic warning. The neural network within HG-DCM allows cross-learning among different pandemics and different locations when fitting the DELPHI model, incorporating data from prior pandemics and metadata to improve incidence curve fitting. This approach preserves the interpretability and epidemiological grounding of the DELPHI model while leveraging historical data through neural network guidance to improve early-stage pandemic forecasting accuracy.

We applied HG-DCM to early COVID-19 forecasting across 227 locations globally, demonstrating that it consistently outperforms the original DELPHI model in early-stage COVID-19 forecasting. This study provides strong evidence that integrating historical data into compartmental models through neural networks can significantly enhance the accuracy and stability of early pandemic forecasting. Furthermore, our comparative analysis reveals that HG-DCM surpasses both state-of-the-art deep learning-based models and compartmental models in early-case forecasting tasks.

### 1.1 Literature Review

Compartmental models have been used to forecast the trend of pandemics since the 20th century [2]. Starting with the simplest SIR (Susceptible, Infectious, Removed) model [32], various compartmental models with different states have shown satisfactory performance in forecasting seasonal pandemics [33, 17]. One of the core strengths of compartmental models is their high interpretability each parameter in a compartmental model usually corresponds to a physical quantity, which provides valuable insights into the pandemic. However, compartmental models also have limitations. Given the inevitable noisiness of the data, compartmental models can significantly overfit during the earliest stage of the pandemic when limited data is available. Furthermore, since compartmental models are inherently modeled for a pandemic in a certain area, it is also not obvious how to incorporate information from other pandemics to augment a compartmental model.

From another direction, machine learning is widely used in time-series forecasting fields such as stock prediction [25], weather forecasting [35], tourism [18], etc. However, most machine learning time-series models are not designed for early-stage pandemic prediction. There are attempts to use advanced deep learning models for pandemic forecasting [31, 30, 36, 9], but these models have been limited to modeling a single pandemic within a single region. Furthermore, these models suffer from the lack of interpretability, which makes the resulting predictions difficult to understand, especially during the early phase of a pandemic.

Overall, there have been few attempts to combine compartmental and deep learning models [16]. Recently, there has been some research that integrates mobility data into the compartmental model through deep learning [8] or utilizes deep learning to estimate the time-varying parameter for the compartmental model [27]. However, these models assume that a significant amount of training data is available for the current pandemic, which makes it unsuitable for early-stage pandemic forecasting.

## 2 Methods

### 2.1 Model Construction

We introduce the History Guided Deep Compartmental Model (HG-DCM), which integrates a deep neural network with a compartmental model to combine the expressivity of a deep learning model and the interpretability of a compartmental model. The model architecture is defined as:

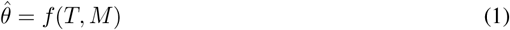

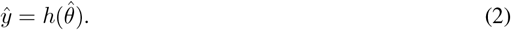

Here *T* and *M* are the time-series and the metadata for the pandemic, which is combined through a deep learning model *f* (·) to create predictions 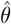 for the parameters for the compartmental model. The final predictions 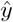 are then calculated by solving an Initial Value Problem (IVP) to map the predicted parameters to a cumulative incidence curve. The key idea is that different pandemics could share a mapping between how the pandemic behaves (*T, M*) and the underlying parameters *θ*, which is captured by the deep learning model *f*. A graphical illustration of the model architecture is showcased in Figure 1. In the following paragraphs, we detail each of the specific structures.

**Figure 1:**
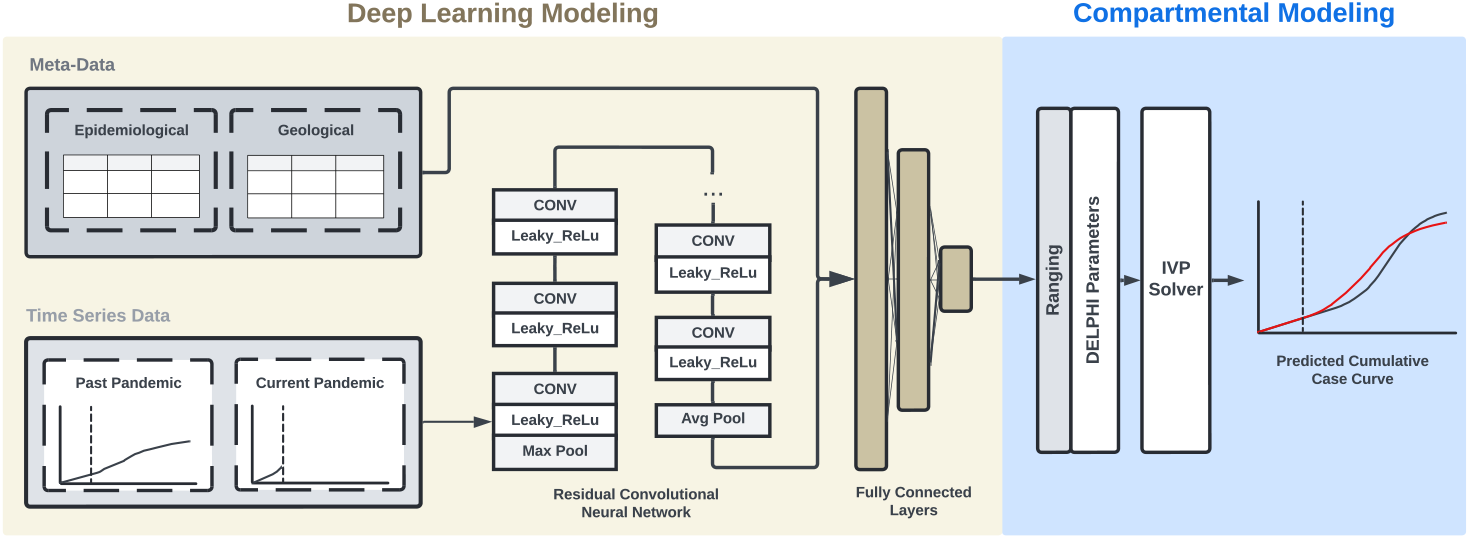
Model Architecture of HG-DCM. HG-DCM consists of two parts: deep learning modeling and compartmental modeling. The deep learning modeling part predicts the compartmental model parameters, and the compartmental modeling section uses the predicted parameters to construct the predicted cumulative case curve for the pandemic.

#### Residual Convolutional Neural Network (RCNN)

We employ a ResNet architecture to train on time-series data from current and past pandemics, where the model *f* (·) predicts pandemic parameters. Since the sample unit is a single pandemic, relying solely on historical data would be insufficient for robust predictions. To address this, we augment historical data using window-shift and masking techniques. For past pandemics, we artificially generate additional samples by applying a sliding window to the training data, predicting future parameters for each windowed segment. The window shifts in 1-day increments up to *M* times or until no future data remains (Figure 2a). For the current pandemic, where future data is unavailable, we apply masking augmentation instead (Figure 2b). To account for variations in case numbers across locations, daily case numbers are log-transformed to enhance model stability. For time series with weekly reporting frequencies or missing data, linear interpolation is used. The ResNet input dimension is [*L, N, D*], where *L* represents the combined lengths of the training and forecasting windows, *N* is the batch size, and *D* is the number of input features (e.g., daily cases, daily deaths). Due to limited data availability, only daily case numbers are used in this study. We also modify the ResNet implementation by removing batch normalization, as differences in batch statistics between past and current pandemics can lead to unstable predictions.

**Figure 2:**
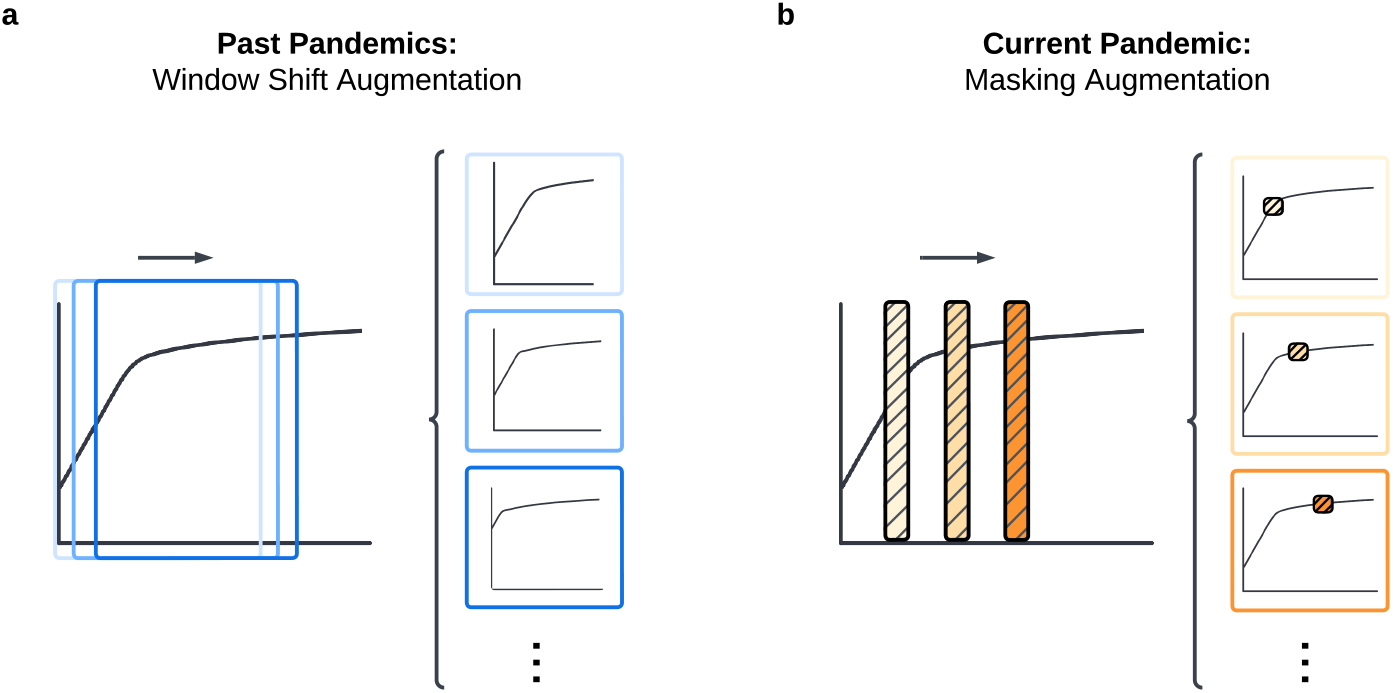
Data Augmentation Methods. **(a)** Window shift data augmentation for past pandemics time series data **(b)** Masking data augmentation for current pandemic time series data

#### Fully Connected Layers

The learned embeddings of the time-series data are concatenated with epidemiological metadata (e.g., transmission pathways) and demographic metadata (e.g., healthcare expenditure). A full table of the metadata is provided in A.1. Meta-data are normalized using min-max normalization and passed through two fully connected layers before concatenating with the time series embeddings output from the RCNN. The concatenated embeddings are passed through fully connected layers to produce parameters for the DELPHI model. To ensure that the produced parameters lie within physical bounds, we utilize a sigmoid ranging function to normalize the predicted parameters between 0 and 1.

#### Compartmental Modeling

We utilize DELPHI [19] as the compartmental model for prediction in this framework. DELPHI is a compartmental epidemiological model that extends the widely used SEIR model to account for under-detection, societal response, and epidemiological trends including changes in mortality rates. The model is governed by a system of ordinary differential equations (ODEs) across 11 states: susceptible (*S*), exposed (*E*), infectious (*I*), undetected cases who will recover (*U*^*R*^) or die (*U*^*D*^), hospitalized cases who will recover (*H*^*R*^) or die (*H*^*D*^), quarantined cases who will recover (*Q*^*R*^) or die (*Q*^*D*^), recovered (*R*) and dead (*D*). The transition rates between the 11 states are defined with 12 parameters, which we predict as 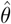 in the HG-DCM framework. To generate the final incidence curve, the estimated parameters are passed through torchODE, a parallel Initial Value Problem (IVP) Solver, to output the predicted cumulative case curve. We used Tsit5 with *a*_*tol*_ = 1 × 10^−8^, *r*_*tol*_ = 1 × 10^−4^ from the torchode package [20] as the ODE solver. We refer the readers to [19] for details on the DELPHI model and its performance.

#### Objective Function

The objective function of HG-DCM is to minimize the loss between the predicted incidence curve and the actual incidence curve of past and current pandemics. The loss of past pandemics includes both the loss of the length-*t* training window and the length-*v* forecasting window (Eqn. 3). The current pandemic loss contains only the training window due to the inaccessibility of the forecasting window in practice (Eqn. 4). Both losses of the past and current pandemics are calculated through a sum of mean absolute error (MAE) and mean absolute percentage error (MAPE) weighted by *α* to balance the effect of the population. The overall loss is calculated by a mean weighted by *β* to balance between past pandemic losses and the current pandemic loss (Eqn. 5). The weight determines the amount of information inherited from past pandemics in predicting the current pandemic. Concretely, the formula for the loss function can be written as:

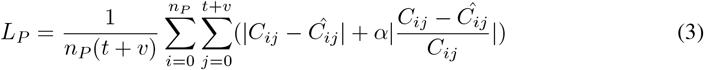

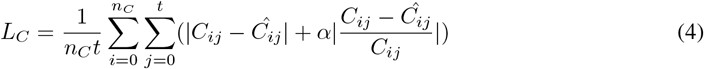

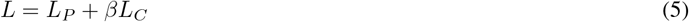

where *n*_*p*_/*n*_*c*_ is the number of samples in the past/current pandemic data, and 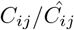 is the actual/predicted cumulative cases of the *i*th pandemic at the *j*th time point.

## 3 Experiments

### 3.1 Experimental Setup

#### 3.1.1 Data

We were unable to find a publicly available database that contained pandemic data from the past. Therefore, we constructed a pandemic dataset, which contains case and death (if available) time series data, pandemic meta-data, and country meta-data for major pandemic outbreaks and seasonal pandemics that have occurred worldwide since 1990. Only pandemics with significant (more than 100) and frequent (daily or weekly) reported incidences are included in the dataset. The dataset includes country-level and domain-level data on the following outbreaks: the 2020 COVID-19 pandemic [6, 5], the 2014 Ebola pandemic [10], the 2003 SARS pandemic [14, 34], the Peru (2000 - 2010) and Puerto Rico (1990 - 2008) Dengue Fever outbreak [21], the 2022 Worldwide Monkeypox outbreak[23], and world-wide seasonal influenza outbreaks (2009-2023) [26, 7].

The time series dataset contains daily or weekly reported cases for each pandemic. The start date of pandemics differs for each location and is set by the first day when the cumulative case number exceeds 100. Epidemiological meta-data with uncertainties that were available at the early stage of the pandemic for each location are collected. The geological meta-data includes 13 country development indicators from the World Bank data [37] for each location in the dataset. The list of meta-data is available in A.1

#### 3.1.2 Setup and Comparison Methods

##### Comparison Methods

We evaluate the model performances on early-stage forecasting tasks, where HG-DCM is used to forecast the cumulative case curve of 12 weeks based on 2/4/6/8 weeks of daily case data. Due to the lack of death data in pandemics prior to COVID-19, only case numbers are used to fit and evaluate the models in the experiments. Locations with no new daily cases reported during the training window are removed from the dataset. The mean and median MAE of the forecasting window between the predicted incidence and the true incidence are used to evaluate model performance. HG-DCM is compared to state-of-the-art compartmental models DELPHI [19] and deep neural network models Gated Recurrent Units (GRU) [4] on the early-stage pandemic forecasting tasks.

##### HG-DCM Setup

Four HG-DCM models are trained using the 2/4/6/8-week training window respectively and predict for 12 weeks. Each HG-DCM is trained separately using the Adam optimizer with a stable learning rate of 1 × 10^−5^. Given the large variation of case numbers among different locations, we use a single-batch training approach, where all samples are passed to the model in one batch. The single-batch approach accelerates the converging by avoiding the turbulent loss curve caused by large variations in incidence numbers among different locations. Dropout or weight decay are not used when training the model. The models used in the comparison are trained for 25k epochs.

##### GRU Setup

We also train a five-layer GRU model for each training window using the same set of past pandemic data as the HG-DCM. We utilize a learning rate of 1 × 10^−3^ as optimized through a grid search. No dropout or weight decay is used.

##### DELPHI Setup

For fitting DELPHI models, the cumulative case curves are fitted separately for each location and each training window. Dual annealing (DA) [38] is used as the optimizer for parameter search. The same default parameter ranges as training HG-DCM are used to fit the DELPHI curve.

### 3.2 Results

#### Early-Stage COVID-19 Forecasting

To assess the effectiveness of integrating deep learning modules into compartmental models, we compare the HG-DCM and DELPHI model in forecasting COVID-19 incidence curves over 12 weeks under different training models. As expected, both models showed improved accuracy with longer training periods. HG-DCM consistently achieved a lower mean MAE across all forecasting tasks (Table 1). Notably, when 8 weeks of data were available, HG-DCM’s 12-week forecasting accuracy was comparable to DELPHI’s in terms of median absolute error (*p >* 0.05). However, with training periods of 6 weeks or less, HG-DCM significantly outperformed DELPHI in median absolute error (*p <* 0.05).For instance, with 6 weeks of training data, HG-DCM is able to reduce median MAE by 21.8% compared to DELPHI, and this improvement increases to 50.1% and 58.7% for 4 and 2-week training windows, respectively. These results underscore the value of integrating historical pandemic information for early-stage pandemic forecasting.

**Table 1:** Model Performance on Covid-19 Early Forecasting. Bold indicates the best performing models (within statistical significance p < 0.05) for each data availability length.

|  |  | 2 Weeks | 4 Weeks | 6 weeks | 8 Weeks |
| --- | --- | --- | --- | --- | --- |
| Mean MAE |  |  |  |  |  |
|  | GRU | **18464.4** | 20832.7 | 24113.0 | 27163.7 |
|  | DELPHI | 611394.9 | 311157.0 | 101547.2 | 21492.6 |
|  | HG-DCM | **18081.6** | **9941.8** | **9302.7** | **5451.6** |
| Median MAE |  |  |  |  |  |
|  | GRU | **4134.5** | 4459.3 | 5360.1 | 5610.3 |
|  | DELPHI | 9258.3 | 4216.6 | 1852.0 | **701.5** |
|  | HG-DCM | **3824.1** | **2103.1** | **1449.3** | **920.3** |

The higher average MAE of the DELPHI model results from overfitting trends within the training window, often leading to overshooting in the cumulative case curve. For example, in the 4-week training task for Switzerland, DELPHI overfitted to the early data, causing a significant overshoot in the 12-week forecast (Figure 3a). By leveraging historical pandemic data, HG-DCM mitigates overfitting and generates forecasts that align more closely with real-world trends.

**Figure 3:**
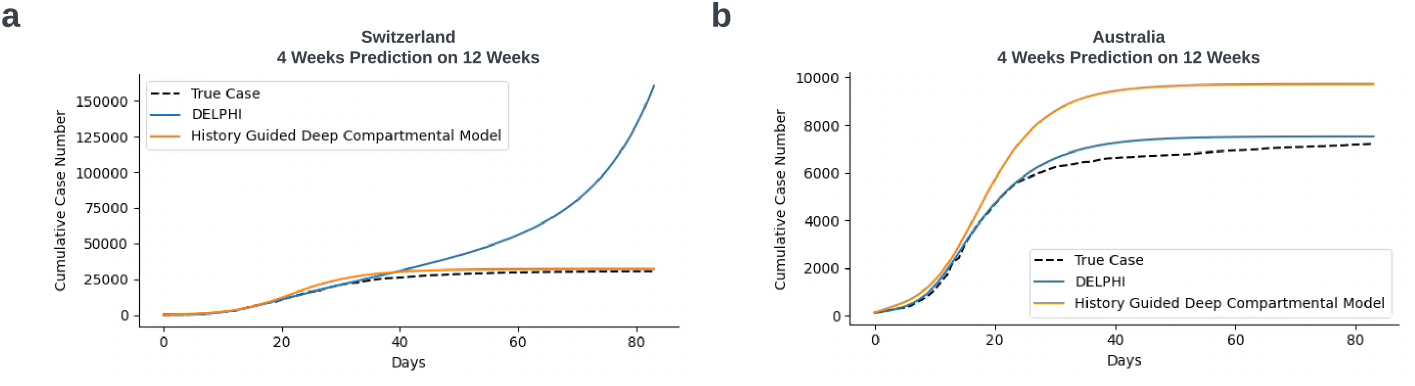
Forecasting Example. **(a)** Sample of a location (Switzerland) where HG-DCM outperforms DELPHI. **(b)** Sample of a location (Australia) where DELPHI outperforms HG-DCM.

To quantify overshooting, we analyzed the distribution of mean absolute errors across locations (Figure 4). DELPHI’s forecasts exhibit long right-hand tails across all training scenarios, reflecting large disparities between predicted and actual curves. In contrast, HG-DCM produces narrower MAE distributions, demonstrating its robustness against extreme forecasting errors. By incorporating guidance from historical pandemics and leveraging information across current locations, HG-DCM effectively reduces overshoot and provides more stable predictions.

**Figure 4:**
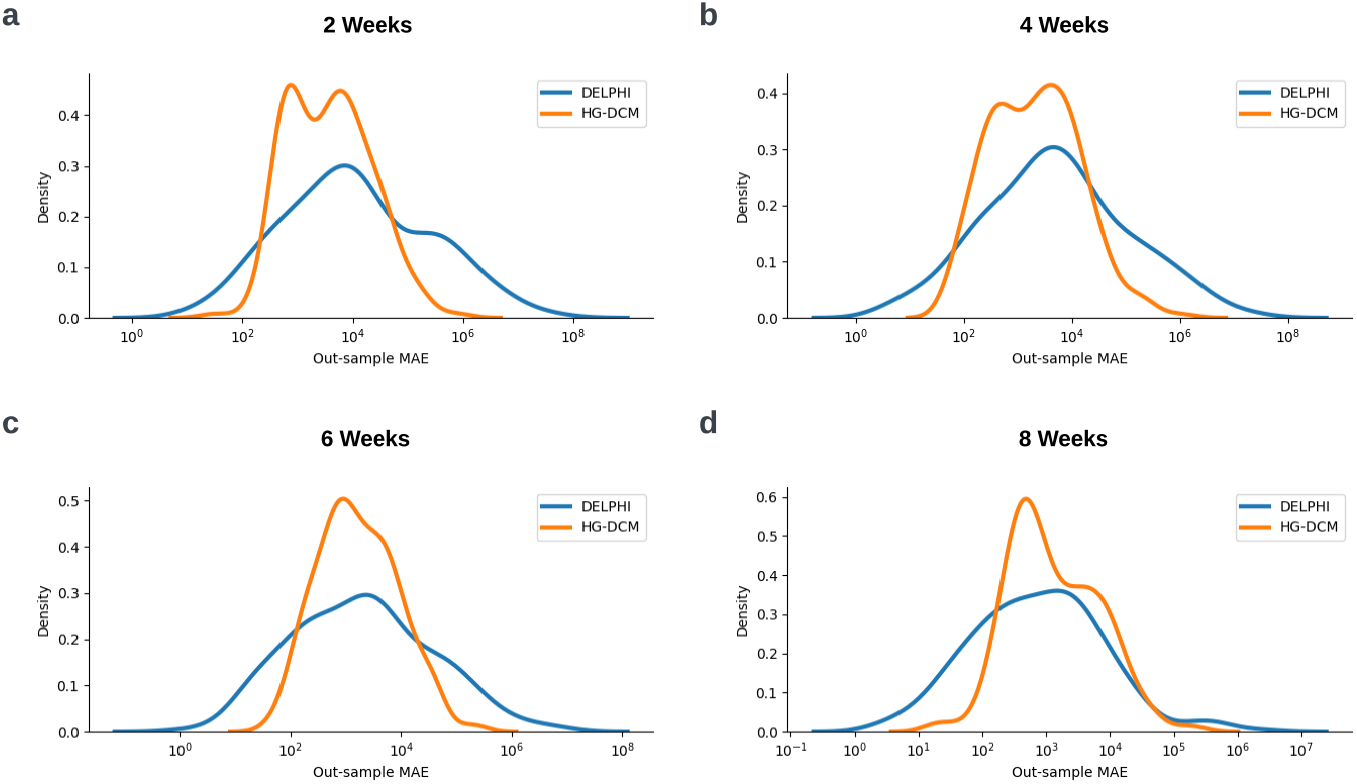
Forecasting Window MAE Distribution. Forecasting window mean absolute error distribution for DELPHI and HG-DCM on COVID-19 12 Weeks Early Forecasting Tasks using **(a)** 2 weeks, **(b)** 4 weeks, **(c)** 6 weeks, and **(d)** 8 weeks of available data.

Furthermore, HG-DCM demonstrates enhanced stability in early-stage pandemic forecasting. Early pandemic data often suffer from inconsistencies due to limited testing, reporting delays, and incomplete case recording. By leveraging epidemiological knowledge from historical pandemics, integrating current pandemic data across locations, and utilizing geospatial meta-data, HG-DCM is better equipped to mitigate the effects of noise. This ensures more reliable and stable estimations, which are critical for timely public health interventions.

HG-DCM also consistently outperforms GRU across the 4-week, 6-week, and 8-week early forecasting tasks (*p <* 0.05), demonstrating significant reductions in median MAE by 83.6%, 73.0%, and 52.8%, respectively. This trend highlights that HG-DCM’s gain does not purely come from the expressivity of a deep learning model - the ability to combine deep learning with an interpretable compartmental model allows HG-DCM to produce significantly better and more physical predictions than a pure deep learning model. We also observe that as training window length increases, the mean and median MAE of GRU increases. We hypothesize this is due to the scale difference among forecasting windows. When the training window is 2 weeks, the forecasting window is from 2 weeks to 12 weeks. For 8 weeks, the forecasting window is from 8 weeks to 12 weeks, which has a significantly higher mean incidence number than that of 2 weeks. The higher mean incidence number in larger training window tasks covers the effect of lowering MAE caused by additional information gained by GRU from extra training window between 2 weeks to 8 weeks, resulting in an increase in mean and median MAE as the training window increases.

Through these experiments, we demonstrated that the HG-DCM provides more accurate and stable forecasting compared to DELPHI at the early stage of the pandemic and the purely deep learning based GRU model. The observation of HG-DCM outperforming DELPHI more on short training windows and GRU more on long training windows shows the importance of integrating epidemiological knowledge from the compartmental model and past pandemic information from deep learning models on early-stage pandemic forecasting tasks.

#### Parameter Inference

One of the key advantages of employing HG-DCM over traditional deep neural networks for pandemic forecasting is its interpretable parameterization. Unlike black-box models, the epidemiologically meaningful parameters predicted by HG-DCM can be extracted before being passed to the Initial Value Problem (IVP) solver, which offers actionable insights.

To illustrate this advantage, we analyzed the parameters inferred by HG-DCM compared to the traditional DELPHI model in an early-stage COVID-19 forecasting task using four weeks of data (Figure 5). The DELPHI model’s parameters exhibited a wide distribution, often leading to unstable forecasts and an overshooting problem. This instability arises because DELPHI fits models independently for each location, amplifying sensitivity to minor noise in the data. In contrast, HG-DCM leverages historical pandemic data and geospatial meta-data, ensuring more robust and consistent parameter estimation.

**Figure 5:**
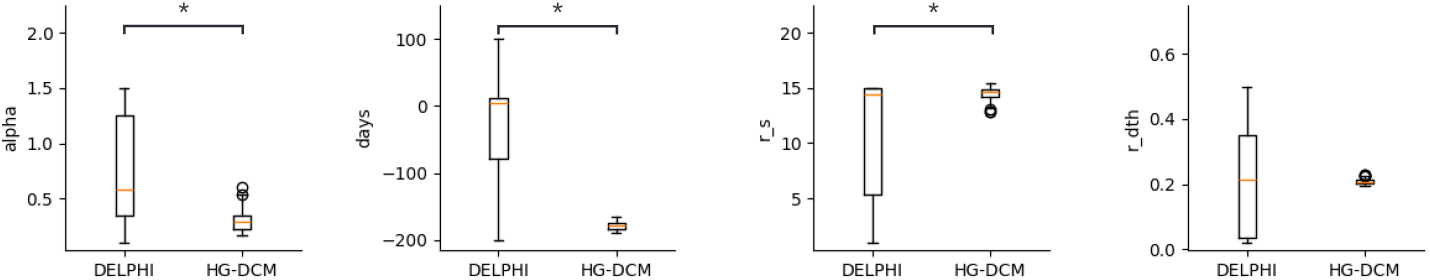
Comparison of fitted parameters in DELPHI and HG-DCM models. The bar graphs show the mean and standard deviation of selected predicted parameters for HG-DCM and DELPHI. The p-values from the Mann-Whitney U test are reported, with p-values *<* 0.05 indicating statistically significant differences.

Statistical analysis using the *Mann-Whitney U Test* [22] confirmed significant differences in key parameters, including the infection rate (*α*), median day of action (*t*_med_), and rate of action (*r*_*s*_), with *p*-values *<* 0.05. Specifically, HG-DCM predicted a lower infection rate and median day of action, while exhibiting a higher rate of action, reflecting its ability to adapt to evolving pandemic dynamics.

Consistency in the rate of death (*r*_dth_) between models further reinforces the reliability of HG-DCM’s parameter estimates. This divergence highlights the complementary strengths of both approaches: DELPHI’s sensitivity to local variation and HG-DCM’s resilience to noise. Together, these insights can facilitate a more nuanced understanding of pandemic behavior. The complete parameter analyses for all 12 DELPHI parameters can be found in Appendix A.2.

#### Model Ablation Study

To understand the contribution of HG-DCM’s design components, we conducted an ablation study by training a Truncated Deep Compartmental Model (T-DCM) that excluded historical pandemic data and meta-data. The T-DCM was trained on datasets with 2/4/6/8 weeks of observations and evaluated on a 12-week forecasting task.

Table 2 shows that T-DCM consistently underperformed HG-DCM across all training window lengths. Notably, HG-DCM achieved significant improvements in both mean and median MAE, with the gap widening as training data length decreased. This result underscores the importance of incorporating historical context and structured meta-data for reliable forecasting in the early stages of pandemics.

**Table 2:** Model performance comparison between HG-DCM and T-DCM.

|  |  | 2 Weeks | 4 Weeks | 6 Weeks | 8 Weeks |
| --- | --- | --- | --- | --- | --- |
| Mean MAE | T-DCM | 18,092.1 | 11,730.5 | 11,982.2 | 8,148.7 |
|  | HG-DCM | **18,081.6** | **9,941.8** | **9,302.7** | **5,451.6** |
| Median MAE | T-DCM | 3,840.1 | 2,450.9 | 2,381.8 | 1,343.9 |
|  | HG-DCM | **3,824.1** | **2,103.1** | **1,449.3** | **920.3** |

#### Early-Stage Monkeypox Forecasting

To demonstrate HG-DCM’s generalizability to future pandemics, we evaluated its performance during the early-stage forecasting of the **Monkeypox (Mpox)** outbreak in Europe and the Americas in 2022 [23]. Using the same methodology as the COVID-19 task, we leveraged historical pandemic data prior to 2022 as guidance.

Table 3 compares HG-DCM’s performance against DELPHI and GRU baselines. HG-DCM achieved the lowest mean MAE across 2-, 4-, and 6-week training windows and the lowest median MAE in most cases, outperforming DELPHI and GRU with statistical significance. These results validate HG-DCM’s ability to adapt to novel pandemics by incorporating historical knowledge while maintaining robustness to noise and data scarcity.

**Table 3:** Model Performance on Mpox Forecasting. Bold indicates the best performing models (within statistical significance *p <* 0.05) for each data availability length.

|  |  | 2 Weeks | 4 Weeks | 6 weeks | 8 Weeks |
| --- | --- | --- | --- | --- | --- |
| Mean MAE | GRU | 1871.1 | 5440.0 | 1776.4 | **2031.6** |
|  | DELPHI | 477973.6 | 393554.2 | 69451.1 | 4181.1 |
|  | HG-DCM | **1516.7** | **1370.6** | **1430.3** | 5441.4 |
| Median MAE | GRU | 1806.8 | 5170.1 | 723.5 | 852.7 |
|  | DELPHI | 1974.3 | **579.3** | 325.2 | **146.7** |
|  | HG-DCM | **1541.2** | 643.9 | **265.4** | **130.6** |

### 3.3 Discussion

In this work, we proposed HG-DCM, a hybrid architecture that bridges compartmental models with deep neural networks for early-stage pandemic forecasting. This framework synergizes the interpretability and domain-grounded rigor of compartmental models with the representational power of deep learning. Specifically, HG-DCM leverages the structured epidemiological insights of compartmental models to ensure plausible predictions while harnessing neural networks to integrate auxiliary information from historical, geographical, and meta-pandemic data. This integration effectively mitigates the pitfalls of overfitting and instability, which often plague individual modeling approaches, particularly during the early phases of a pandemic when data is sparse and noisy.

Our results demonstrate that HG-DCM outperforms both standalone compartmental models and purely deep learning-based models on early forecasting tasks. By offering a more robust and accurate early-stage estimation of pandemic trends, HG-DCM addresses critical challenges in public health response, such as resource allocation and policy planning. In particular, its ability to produce stable, noise-robust predictions reduces erratic shifts in trend forecasts, enabling more confident decision-making and minimizing the risks of resource misallocation caused by extreme over- or underestimation.

A key strength of HG-DCM is its interpretability, which remains a cornerstone for pandemic forecasting applications. While deep learning methods often function as opaque black boxes, HG-DCM retains the parameter-driven transparency of traditional compartmental models, with fitted parameters offering actionable epidemiological insights. For instance, in our implementation with the DELPHI compartmental model, the extracted parameters maintain clinical relevance, providing healthcare providers with early and interpretable guidance on the potential trajectory of a pandemic. This interpretability is invaluable for building trust with stakeholders and ensuring actionable insights.

Beyond its strong predictive performance and interpretability, HG-DCM exhibits significant architectural flexibility. In our experiments, we employed a ResNet-based module for temporal representation learning and the DELPHI model for cumulative case curve estimation, selected based on empirical evaluations. However, the modular design of HG-DCM allows for seamless integration of more advanced compartmental models or neural network architectures as they emerge. This adaptability positions HG-DCM as a forward-compatible framework capable of evolving alongside advances in epidemiology and machine learning.

In summary, HG-DCM provides a practical, interpretable, and extensible solution for early-stage pandemic forecasting. By demonstrating the utility of combining epidemiological and deep learning methodologies, our work highlights the potential of hybrid approaches to address complex forecasting challenges in the face of limited data and high uncertainty. Future research may explore augmenting HG-DCM with additional data modalities, enhancing its generalizability to a broader spectrum of infectious diseases, and extending its application to real-time adaptive forecasting.

## 4 Limitation

While HG-DCM demonstrates strong performance in early-stage pandemic forecasting, it is not without limitations. One notable challenge lies in handling the high variability of incidence rates across different geographical regions. This variability renders conventional normalization techniques, such as batch normalization, unsuitable for the stable estimation of model parameters. Empirical experiments revealed that incorporating batch normalization resulted in unstable predictions, while layer normalization caused critical information loss, impeding model convergence. As a result, no normalization technique was employed in HG-DCM, which, while stabilizing predictions, increased the overall training time due to slower convergence.

Another limitation relates to the availability and quality of historical pandemic data. The COVID-19 pandemic provided the first instance of high-resolution, daily time series data, which proved instrumental in enabling robust model training and evaluation. In contrast, earlier pandemics, such as Ebola, SARS, and Dengue Fever, often lack comparable data granularity. These datasets are frequently reported in weekly aggregates, requiring interpolation to align with HG-DCM’s daily prediction framework. Linear interpolation, while a practical workaround, introduces approximation errors, particularly during the critical early stages of a pandemic when precise trend estimation is most needed. This limitation highlights the dependency of HG-DCM on the quality and resolution of input data, which directly impacts its forecasting accuracy.

Furthermore, while the COVID-19 pandemic raised awareness of the importance of robust pandemic data collection, the availability of high-quality, real-time data remains inconsistent across regions and diseases. Recent outbreaks, such as Monkeypox in 2022, demonstrate progress in this area, with improved public access to daily incidence and mortality data. However, disparities in data quality and completeness persist globally, posing ongoing challenges for comprehensive model training.

Despite these constraints, HG-DCM’s modular and flexible design ensures its applicability to evolving data landscapes. As the availability and fidelity of historical pandemic datasets improve, future iterations of HG-DCM can leverage these advancements to further enhance its capabilities. Addressing the aforementioned limitations will be critical for developing more generalizable and efficient forecasting frameworks for infectious disease outbreaks.

## 5 Conclusion

In this work, we introduced HG-DCM, a novel deep compartmental architecture designed to enhance early-stage pandemic forecasting. Our approach integrates historical pandemic data and metadata through a deep learning framework coupled with a compartmental modeling component that generates interpretable forecasts. We demonstrated that HG-DCM outperforms both traditional compartmental models and standalone deep learning models in early-stage forecasting tasks.

These results highlight the promise of deep compartmental models for pandemic forecasting and underscore the value of incorporating historical pandemic data. Future work could focus on integrating additional data sources, such as mobility patterns, policy interventions, or other metadata, to further improve forecasting accuracy. Moreover, adapting HG-DCM for real-time applications represents an exciting avenue for research. We believe this work establishes a foundation for leveraging past pandemics through deep learning to inform future forecasting efforts.

## Data Availability

All data produced are available online at https://github.com/AlexWei21/Pandemic-Database

## A Appendix

### A.1 Meta-Data

To incorporate epidemiological and geographical information into early-stage pandemic forecasting, we collected 7 epidemiological and 13 geographical metadata for each location and pandemic. Data unavailable during the early stages were marked as missing in the dataset. A detailed list of the metadata collected is provided in Table A.1.

**Table A.1:** Meta data table for training HG-DCM. The meta-data table including the epidemiological metadata and geological metadata used in training HG-DCM

| **Epidemiological Meta Data** | **Geological Meta Data** |
| --- | --- |
| Basic Reproduction Rate ($R_0$) | Population |
| Transmission Pathways | Net lending/borrowing |
| Mortality Rate | Current Health Expenditure per capita |
| Average Hospitalization Length | Population Density |
| Hospitalization Rate | GNI per capita |
| Latent Period | GDP per capita |
| Incubation Period | Physician per 1,000 people |
|  | Urban Population Living in Slums |
|  | GDP |
|  | External Health Expenditure per capita |
|  | Air Transport |
|  | Government Effectiveness |
|  | Domestic General Government Health Expenditure per Capita |

### A.2 COVID-19 Early-Stage Forecasting Parameter Analysis

To better interpret the predictions, we analyzed the parameters inferred from HG-DCM compared to DELPHI in an early-stage COVID-19 forecasting task using four weeks of data. Among the 12 parameters, predicted infection rate (*α*), median day of action (*days*), rate of action (*r*_*s*), Initial Infection (*k*_2_), Median day of jump (*t*_*jump*), rate of case resurgence (*std*_*normal*), and *k*3 are significantly different between DELPHI and HG-DCM. HG-DCM model tends to predict a lower *α, days, t*_*jump*, and *std*_*normal*, whereas predicting a higher *r*_*s, k*2, and *k*3. The divergent prediction set of parameters from two different forecasting methods provides a more comprehensive understanding of the pandemic. Additionally, DELPHI and HG-DCM produced consistent predictions for the rate of death (*r*_*dth*), initial mortality rate (*p*_*dth*), rate of mortality decay (*r*_*dthdecay*), Initial Exposure (*k*1), and magnitude of the jump (*jump*), reinforcing the validity of the inferred parameters (Figure A.1).

### A.3 Forecasting Window MAE Distribution of Early-Stage Monkeypox Forecasting

To visualize the performance of HG-DCM and DELPHI in early-stage Monkeypox forecasting, we plot the distribution of Mean Absolute Error (MAE) values for the predictions. For the 2-, 4-, and 6-week forecasting tasks, HG-DCM predictions exhibit a narrower MAE distribution compared to DELPHI, aligning with similar observations from early-stage COVID-19 forecasting tasks. The overlapping prediction distributions of HG-DCM and DELPHI for the 8-week forecasting task suggest that HG-DCM can achieve comparable performance to DELPHI when sufficient information is available for current pandemic forecasting (Figure A.2).

### A.4 HG-DCM Outputs More Stable Forecasting than DELPHI

In the early stages of the pandemic, even minor fluctuations in a single data point can significantly impact the trend when fitting traditional compartmental models. For example, DELPHI’s forecast for the cumulative COVID-19 case curve in the United States varied substantially depending on whether it used 4, 6, or 8 weeks of data (Figure A.3). Specifically, the overshootings observed with 4- and 8-week training window prediction tasks are largely attributable to isolated increases in daily case counts at the end of these periods, likely due to data noise. DELPHI overfits the noise in the data and produces an overshooting curve for the 12-week forecasting. In contrast, HG-DCM produced consistent prediction curves across the 4-, 6-, and 8-week training windows, demonstrating its robustness against data noise.

**Figure A.1:**
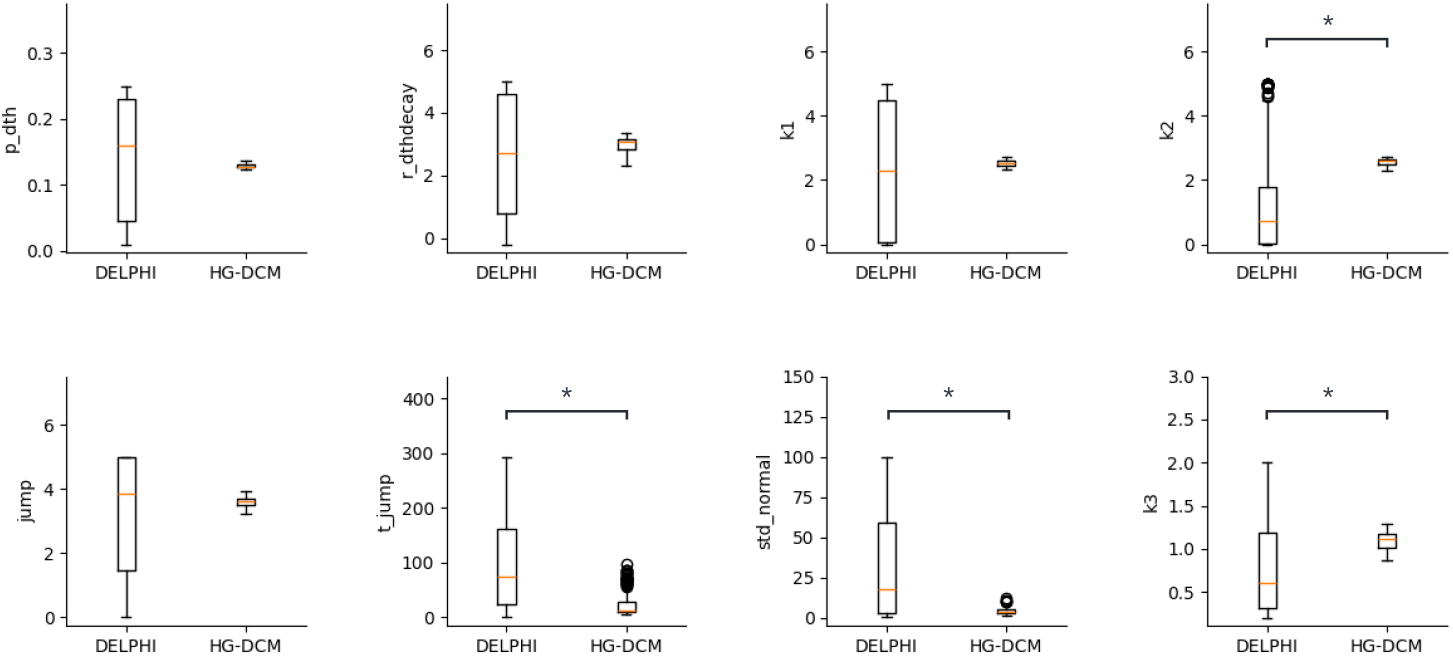
Comparison of fitted parameters in traditional DELPHI model and HG-DCM. The 8 bar graphs show the mean and standard deviation of the remaining 8 predicted parameters from two different approaches that are not shown in the main text. Mann-Whitney U test is used to calculate the p-value of the two groups. Pairs with p-values < 0.05 are considered significantly different.

**Figure A.2:**
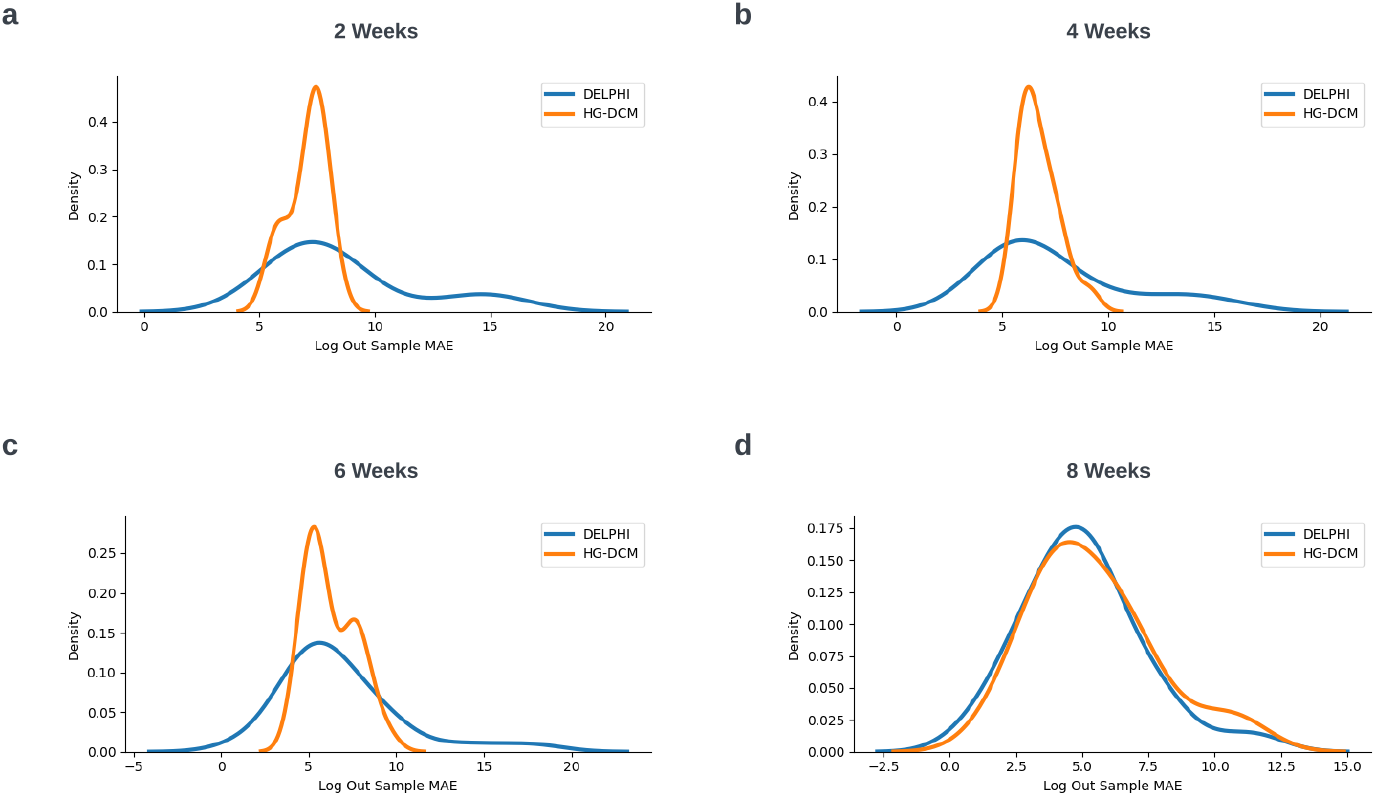
Forecasting Window MAE Distribution of Mpox Early Forecasting Tasks. MAE distribution of DELPHI and HG-DCM on 12-Week Early-Stage Monkeypox Forecasting Tasks using **(a)** 2 weeks, **(b)** 4 weeks, **(c)** 6 weeks, and **(d)** 8 weeks of available data.

**Figure A.3:**
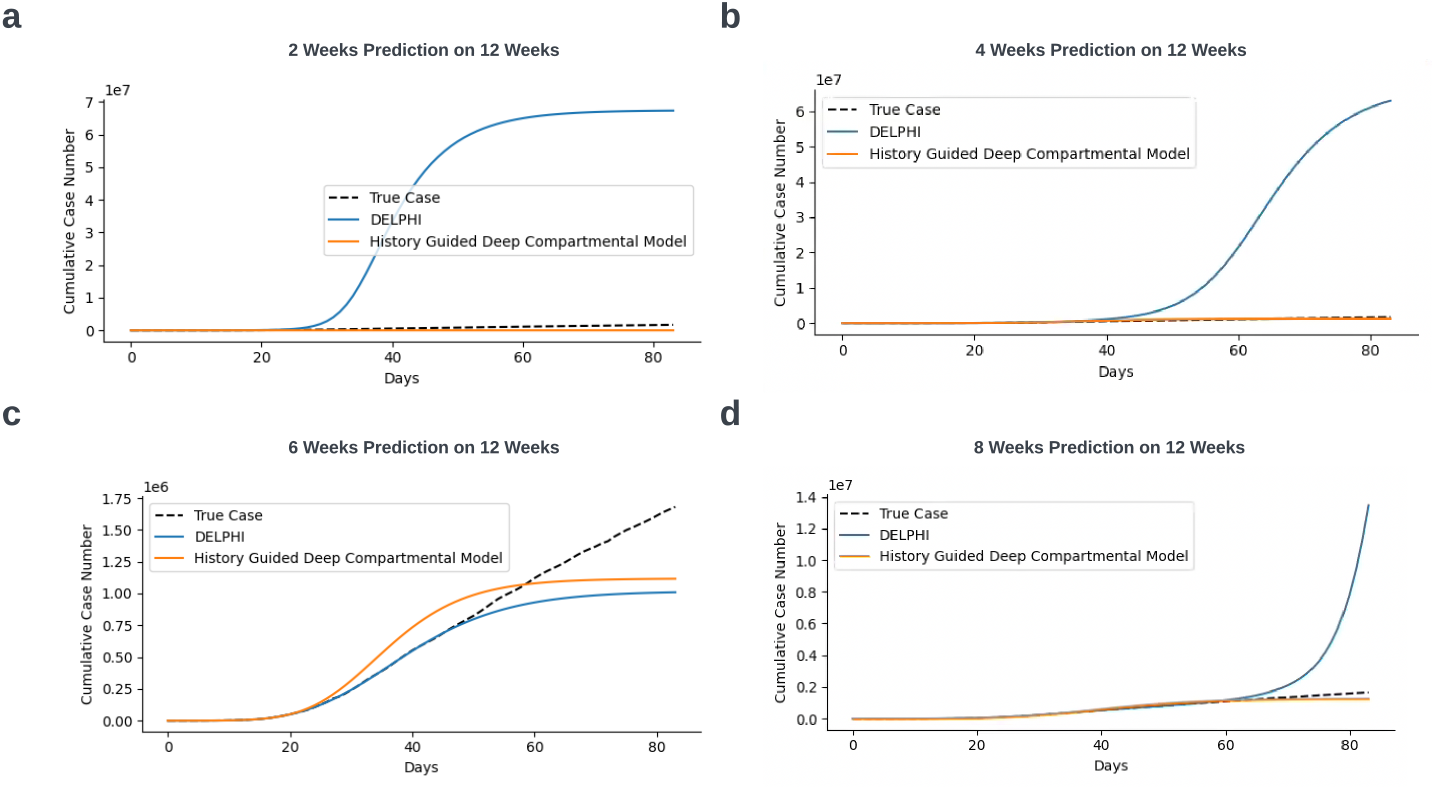
12-Week Forecasting of United States Incidence Curves by HG-DCM and DELPHI. The 12-week forecasting produced by HG-DCM and DELPHI using **(a)** 2 weeks, **(b)** 4 weeks, **(c)** 6 weeks, and **(d)** 8 weeks of available data for the United States.

